# Advanced Practice Physiotherapists succeed in Shared Decision-making: A Mixed-Methods Study of 5123 Patients with Musculoskeletal Pain

**DOI:** 10.64898/2025.12.20.25342750

**Authors:** Joanne H Thompson, Gareth Whelan, Glyn Elwyn, Kristian Damgaard Lyng

**Author notes:** **Corresponding author:** Kristian Damgaard Lyng, Department of Health Science and Technology, Aalborg University, Selma Lagerlöfs Vej 249, 9260 Gistrup, Denmark.

## Abstract

**Question:** Do advanced practice physiotherapists succeed in conducting shared decision-making (SDM)?

**Design:** A prospective cross-sectional observational study of patients with musculoskeletal pain and their perceptions of SDM measured using the CollaboRATE instrument. Written feedback was collected after consultation to gain understanding of patients’ experiences of their consultations.

**Participants:** Chronic pain patients (n = 5123) diagnosed with either upper limb (n = 1230, 24%), lower limb ((n = 3044, 59.4%), or foot/ankle pain (n = 849, 16.6%) consulting an advanced practice physiotherapist across two hospitals in the UK between January 2023 and December 2024.

**Results:** Mean total CollaboRATE scores across all items were 11.9 (±0.53). Overall, 4906 (95.8%) of participants gave the maximum CollaboRATE score of 12. No significant differences were observed between sites (p < .001). A regression model including site and pain category was statistically significant but explained minimal variance (R² = 0.004), suggesting other factors contribute more substantially to SDM perceptions. From 949 patient responses, our qualitative analysis revealed a generally positive SDM experience with advanced practice physiotherapists. Feedback clustered around five key themes: 1) feeling valued and involved, 2) communication, expertise, and clarity, 3) compassionate and professional care, 4) efficient organisation, and 5) negative experiences.

**Conclusion:** Advanced practice physiotherapists were largely successful in facilitating SDM, with patients reporting positive experiences across both clinical sites. While quantitative findings showed minimal influence of site or pain category, qualitative insights highlighted the importance of clinician communication, empathy, and involvement of patients in care planning as key drivers of SDM perceptions.

## Background

Musculoskeletal pain disorders represent the leading cause of disability worldwide, significantly impacting individuals’ quality of life and placing a considerable burden on healthcare systems ^1,2^. Effective management of musculoskeletal pain relies on a combination of clinical expertise, patient preferences, and evidence-based practices ^3^. To align clinical expertise and patient preferences with evidence-based practices, shared decision-making (SDM) is often proposed as an effective approach to ensure that patients actively participate in decisions about their care ^4^. Using SDM provides a positive shift in patient autonomy and decision making, enabling people to be informed and to express ‘what matters’ to them ^4,5^. While the benefits of SDM are well-documented, its implementation varies widely across different healthcare settings within musculoskeletal pain ^6,7^. While research on SDM in musculoskeletal care is growing, the field has largely been focusing on physician-led consultations rather than physiotherapist-led care ^8^. Advanced practice physiotherapists are increasingly recognized for their ability to lead frontline musculoskeletal care, often operating with extended scopes of practice and in autonomous roles ^9,10^. Their role offers an opportunity to implement SDM principles in day-to-day care, potentially improving patient outcomes and streamlining resource use ^11^. While SDM should be applicable to all patients, regardless of background, it is a concern whether socioeconomically vulnerable patients with low levels of education and income experience the same quality of care in relation to SDM ^12^. Little is known about how socioeconomic factors influence SDM in these settings, but a systematic review found that SDM interventions significantly improve outcomes for disadvantaged people ^13^. However, there is a significant lack of knowledge regarding the impact of advanced practice physiotherapists on SDM and how socioeconomic status influences this dynamic. Therefore, this study aimed to examine the extent of patient-perceived success towards SDM during musculoskeletal care consultations with advanced practice physiotherapists across two clinical settings with differing socioeconomic profiles. Additionally, it sought to compare CollaboRATE scores between the two sites and lastly, the study aimed to identify key themes related to patients’ experiences with advanced practice physiotherapists.

## Methods

### Study design

This study was designed as a prospective cross-sectional study. The study was registered and approved by the Clinical Governance team at York and Scarborough Teaching Hospitals NHS Trust with no requirement for ethics approval. The study was conducted as part of a service evaluation across two clinical sites, which did not permit the collection of identifiable data such as name, age, or gender. Data collection of the study took place between January 2023 to December 2024. The reporting of the study adhered to The Strengthening the Reporting of Observational Studies in Epidemiology (STROBE) checklist ^14,15^.

### Settings

The cross-sectional study was carried out prospectively at two locations: Clifton Park Clinic located in York, UK and The New Selby War Memorial Hospital located in Selby, UK. Both clinics form part of the Musculoskeletal Service delivered by York and Scarborough Teaching Hospitals NHS Foundation Trust. The services provide comprehensive musculoskeletal healthcare to an estimated population of approximately 350,000 individuals. This service is delivered through two distinct teams. The musculoskeletal physiotherapy teams offer support, advice, and physiotherapy treatment for patients experiencing new or recurrent musculoskeletal problems, as well as those managing long-term musculoskeletal conditions. Complementing this, the musculoskeletal advanced practice team supports patients along established outpatient diagnostic pathways to determine the correct treatment and management options available to them and acts as an interface to secondary care orthopaedic services. Healthcare delivery within the service is concentrated in the two major population centres of York and Selby. York, home to over 202,821 residents according to the 2021 UK Census report, serves as a regional economic hub with a diversified economy spanning biotechnology, digital industries, financial services, tourism, and education ^16^. The city also hosts two universities and benefits from a generally affluent population characterized by higher life expectancies, lower obesity and smoking rates, and a smaller proportion of individuals residing in areas within the lowest 20% of the Index of Multiple Deprivation. In contrast, Selby, with a smaller population of approximately 92,000, has historically relied on manufacturing, mining, and agriculture as its economic backbone ^16^. While these sectors remain influential, Selby retains a strong manufacturing and agricultural presence. The population of York is generally more affluent than that off Selby, have higher life expectancies, lower rates of obesity and smoking, and a smaller percentage of the population within the lower 20% of Index of Multiple Deprivation ^16^.

### Participants

#### Inclusion

Patients had either been referred from their primary care health provider or self-referred to the service and had been triaged by an advanced care physiotherapist as needing an appointment with the advanced practice team. The criteria for triage to the advanced practice team were:

- Traumatic injury >6 weeks who no longer interact with an emergency care pathway and there is ongoing significant sequelae.
- Patients who have not responded to first-line MSK treatment AND where there is a possible interventional treatment available (e.g. OA joint, degenerative rotator cuff tear etc)
- Patients where there is diagnostic uncertainty, and an advanced practice physiotherapist’s opinion is explicitly sought, including appropriate requests for consideration of diagnostic imaging.
- Appropriate requests to access ultrasound-guided injection

#### Variables and data measurements

The primary outcome variable for this study is patient-perceived SDM, measured using the CollaboRATE instrument ^17^. CollaboRATE is a brief, patient-reported measure specifically designed to evaluate the quality of SDM during clinical encounters ^17^. It assesses patients’ perceptions of how well clinicians involve them in decisions about their care, focusing on three core domains: explaining options, listening to patients, and ensuring decisions reflect patient preferences ^17^ and is validated to assesses the extent to which patients feel involved in decision-making during their consultation ^18,19^. Each question is scored on a 5-point anchored scale from 0 (no effort was made), 1 (a little effort was made), 2 (some effort was made), 3 (a lot of effort was made), and 4 (every effort was made). A total score is based on the sum of the scores of the three items with a lower score indicating a patient perceived lower quality of SDM during their clinical consultation ^17^. To accommodate the ceiling effects often exhibited by the CollaboRATE measure, we conducted a top-score analysis ^20^. Therefore, we additionally reported the number and proportion of patients who achieved the highest possible score (top score = 12). The main independent variables included the clinical setting, categorized as York or Selby, and the pain condition, classified into upper limb (including cervical spine), lower limb (including lumbar spine), or foot and ankle pain. The date of consultation is included as a continuous variable to account for any temporal trends. Additionally, qualitative data in the form of free-text patient comments were collected to provide further insights into their experiences with the advanced care physiotherapist. All new patients attending a first appointment with an advanced care physiotherapists at the two locations were offered the opportunity to complete the CollaboRATE instrument by the administrative team at the clinic’s reception. The CollaboRATE data were collected anonymous and immediately following each consultation.

#### Statistical analysis

The primary outcome variable is the total CollaboRATE score. The distribution of total CollaboRATE scores were assessed for normality using descriptive statistics and visualizations on Q-Q plots. Descriptive statistics was used to summarize all variables, with means and standard deviations or medians and interquartile ranges for continuous data and counts and percentages for categorical data. Due to known ceiling effects in patient-reported experience measures, a top-score analysis (i.e., the proportion of patients achieving the maximum total score) was determined a priori to enhance the detection of variation. To compare CollaboRATE scores between the two sites, an independent samples t-test or Mann-Whitney U test was be applied, while a one-way ANOVA or Kruskal-Wallis test was be used for differences across the three pain-condition categories. Furthermore, we conducted a multivariable linear regression analysis to examine whether clinical site and pain category significantly predicted variation in CollaboRATE scores ^21^. The regression model included total CollaboRATE score as the dependent variable and site and pain category as independent predictors. Statistical significance was evaluated at *p* ≥ 0.05, and effect sizes were assessed using standardized beta coefficients (β). Multicollinearity among predictors was assessed using Variance Inflation Factor (VIF) and condition index, with VIF values close to 1.0 and a condition index below 10 indicating a negligible collinearity ^22^.

#### Thematic analysis

The qualitative feedback collected from the patients were analysed via Braun and Clark’s reflective thematic analysis approach ^23^. The feedback was analysed by two authors (JT and KDL) in collaboration with at least one member of the research group (GW or GE). Qualitative data were analysed through a stepwise process of familiarization, coding, theme development, and reflective discussion, guided by an inductive approach. The lead analyst conducted in-depth familiarization with the written feedback and then coded the data using NVivo 12 to categorize and connect emerging ideas. Preliminary insights were shared with the entire author group, and semantic themes were refined iteratively by merging and reorganizing related codes. Any overlaps or uncertainties were resolved collaboratively with the senior author (KDL), ensuring a nuanced interpretation of the data. Through repeated cycles of coding and reflection, broader latent themes were identified and named. This final set of themes formed the basis of the narrative synthesis presented in the results section. In addition to the thematic analysis, we conducted a sentiment analysis to assess the emotional tone of the feedback and complement the interpretive depth with an overview of overall patient sentiment.

## Results

Between January 2023 and December 2024, 5123 patients were recruited from Clifton Park Clinic, York (n = 4352, 85%) and Selby New War Memorial Hospital (n = 771, 15%). The majority were diagnosed with lower-limb pain (n = 3044, 59.4%), followed by upper-limb (n = 1230, 24%) and foot-and-ankle pain (n = 849, 16.6%) conditions. The mean (SD) CollaboRATE scores for items 1, 2, and 3 were 3.97 (0.19), 3.96 (0.20), and 3.96 (0.20), respectively, with a total summed score of 11.9 (0.53), indicating favourable SDM perceptions. Overall, 4906 patients (95.8%) gave the maximum possible total score of 12, (i.e., reflecting a strong ceiling effect). The proportion of top scores was 96.2% (n = 2929) of the 3044 patients with lower-limb pain, 95.4% (n = 1173) of the 1230 patients with for upper-limb pain, and 94.7% (n = 804) of the 849 patients living with foot-and-ankle pain conditions. When comparing locations, 4180 of 4352 patients (96.0%) at Clifton Park Clinic and 726 of 771 patients (94.2%) at Selby New War Memorial Hospital achieved top scores.

An independent samples t-test found no statistically significant difference in CollaboRATE scores between York (M = 11.9, SD = 0.50) and Selby (M = 11.8, SD = 0.71), t(907) = 1.891, p < .001, suggesting similar SDM perceptions across sites. A one-way ANOVA revealed a small but significant effect of pain category on CollaboRATE scores, F(2, 5120) = 6.899, p = .001, η² = .003. Post-hoc Tukey’s HSD test showed that patients with lower-limb pain (M = 11.93, SD = 0.39) reported significantly higher SDM perceptions than those with foot & ankle pain (M = 11.85, SD = 0.76, p = .002, 95% CI [0.0225, 0.1205]). No significant differences were found between upper-limb and lower-limb pain (p = .062) or upper-limb and foot-and-ankle pain (p = .415).

A multiple linear regression examining site and pain category as predictors of CollaboRATE scores was statistically significant, F(2, 5120) = 10.138, p < .001, but explained minimal variance (R² = 0.004). Pain category (β = -0.038, p < .001, β = -0.053) and site (β = -0.054, p = .011, β = -0.036) were both statistically significant predictors, but effect sizes were small. Collinearity diagnostics (VIF ≈ 1.001, Condition Index < 10) confirmed that predictors contributed independently. Overall, while site and pain category significantly predicted SDM perceptions, their low explanatory power suggests that other factors may play a more substantial role in shaping patient-reported SDM experiences.

### Patient perspectives following consultations

Patient feedback highlighted a range of mostly positive experiences, centred on themes of feeling valued and involved, receiving compassionate and professional care, experiencing effective communication, expertise, and clarity, and dealing with an efficient organisation. A smaller subset of patients described negative experiences, revealing tensions between expectations, delivery, and organisational barriers. Across these themes, patients emphasized the importance of being heard, respected, and included, which supported an increase in sense of trust and satisfaction with the consultation. The themes can be seen visualized in **figure 1**.

**Figure 1.**
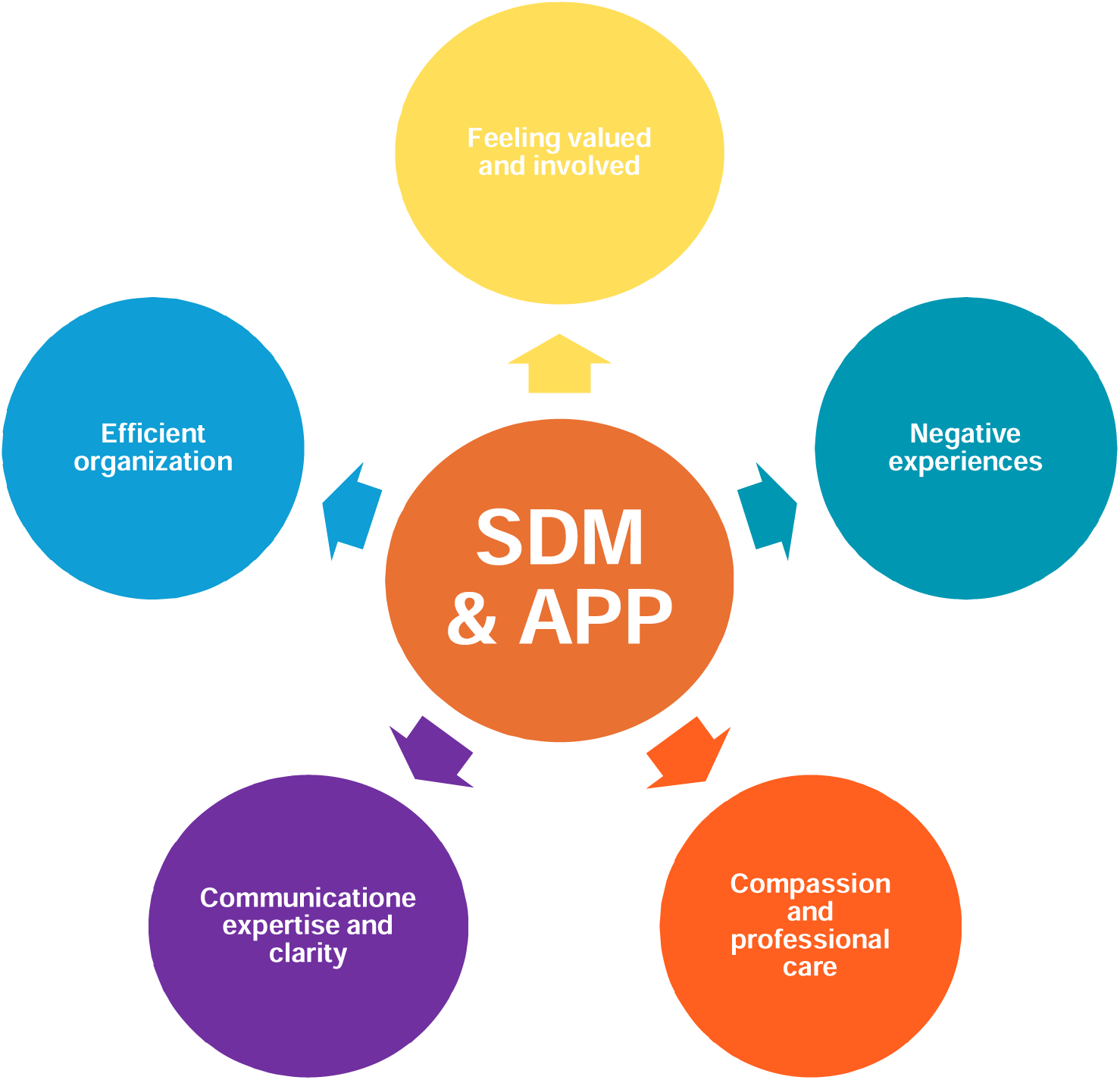
Overview of themes.

#### Feeling valued and involved

Many patients expressed a sense of being respected and the feeling of being involved in decision-making, with one participant stating, “He (*the physiotherapist) was excellent, thorough, and (he) involved me in decision making (ID 2517)*”. Another noted how this involvement increased the patients’ beliefs in themselves and their ability to self-manage “*She (the physiotherapist) made me feel reassured, and she was very good at explaining my condition. I now feel confident in self-managing my condition (ID 3077)*”. Additionally, several patients described their feelings and needs being valued in their consultation including “*it was lovely to feel valued, that my worries were addressed, and that the ball is now rolling (ID 79)*”, “*all the staff were super. My physiotherapist made feel that my health care was being manged with me rather than done to me (ID 328)*” and “*I’m very happy with advice given, clear about my plan moving forward, listened to me and I felt he understood my needs (ID 571)*”. This perception of being included and valued helped foster a sense of shared responsibility in care planning and decision-making.

#### Communication, expertise, and clarity

A recurring theme was the value of clear communication, with one patient noting, “I’m *very positive of the consultation, she (the physiotherapist) fully explained my condition, she was clear and to the point in both advantages and limitations of the procedure and honesty about timelines of my condition (ID 1083)*”. Patients also frequently emphasized thorough assessments and the clarity of diagnoses, with comments such as, “*No one I have seen before has been as thorough or caring. He (the physiotherapist) is a real asset to the team (ID 1659)*”, “*She (the physiotherapist) was really good indeed, she gave a well explained diagnosis and provided me with future choices that were very well explained as well (ID 1866)*”, and “*Examination was very thorough and made me feel that a diagnosis might be found, thank you (ID 202)*”. Along this theme, most patients praised the physiotherapist’s knowledge and competencies as a key driver for positive reviews including “*He (the physiotherapist) was professional, personable, knowledgeable, and very thorough. Listened at every stage and assessment was excellent (ID 104)*”.

#### Compassionate and professional care

Compassion appeared to facilitate trust in both the physiotherapist and the system, which possibly facilitated positive experiences and outcomes. Many patients praised the clinicians’ ability to combine empathy with professionalism, describing how this balance made them feel both emotionally supported and clinically safe. This foundation enabled more open conversations and joint planning. One patient explained, “*She (the physiotherapist) had a combination of professionalism and empathy which was perfect. Information was clear and covered everything efficiently and in a kindly manner (ID 1311)*” and another explained how the compassionate and professional care made them feel safe “*My physiotherapist was very approachable, didn’t feel judged and I felt safe, this is a huge positive for me (ID 1698)*”.

#### Efficient organization

A well-structured and timely consultation was described as contributing to a sense of control and confidence in the process. One patient highlighted this in the following “*She (the physiotherapist) was very friendly, understanding, identified the problem, discussed further investigation and possible treatment, she showed me my x-ray, she had already reviewed my history, which is very complex. She was really prepared (ID 1812)*”. Several participants praised the high-quality care and the efficiency with the organization was seen as a facilitator for SDM, with one patient noting “*I was very impressed with my appointment, everyone was efficient, professional physio, thorough, and informative for the future plan (ID 2368)*”.

#### Negative experiences

However, while most comments were positive or neutral, some concerns were raised about time constraints during consultations and the organisational context. A small number of patients mentioned feeling that their appointments were rushed, with one stating, “*He (the physiotherapist) felt rushed like he did not have enough time (ID 15)*”, which possibly negatively affected SDM. Lastly, several participants noted challenges in finding the right location and delays, “*very hard to find form the main roads, this is not very good, and the signposting are very bad (ID 3453)*” and “*the staff was good, but only niggle was a 30-minute wait for my appointment (ID 3459)*”. These comments suggest that while most patients felt valued and engaged in SDM, ensuring adequate time for consultations, and addressing potential organisational issues remains of importance shaping the entire experience.

## Discussion

This study found high patient-perceived SDM across both clinical sites, with minimal differences between York and Selby. While lower-limb pain patients reported slightly higher SDM perceptions than those with foot and ankle pain, effect sizes were small, and regression analysis showed site and pain category explained only 0.4% of variance, suggesting other factors play a more significant role. This was further reflected in the top-score analysis, where 95.8% of patients gave the highest possible CollaboRATE score, indicating a strong ceiling effect. Qualitative feedback supported these findings, highlighting the importance of feeling valued, clear communication, clinician expertise, and compassionate care in shaping patients’ experience of SDM. Overall, clinician-patient interaction appears to be the strongest driver of SDM perceptions, outweighing demographic, or clinical factors.

## Implications for physiotherapy practice

This study highlights that those patients attending advanced practice physiotherapy services report consistently high perceptions of SDM, regardless of clinical site or presenting condition. The findings suggest that the quality of clinician–patient interaction, particularly clear communication, active listening, and structured explanation of care options, is the primary driver of perceived SDM. This reinforces the critical role of relational skills in physiotherapy practice, especially in first contact and extended-scope roles. While SDM is a recognised element of patient-centred care and is embedded in physiotherapy societies internationally ^24,25^, its routine implementation remains a clinician-perceived barrier in time-limited consultations ^11,26^. Importantly, newer research has raised attention to the uncertainties related to time requirements of SDM, emphasising that SDM do not necessarily requires more time ^27,28^. The perceived mismatch between the time required and the time available for SDM highlights the need for targeted training, either through continuing professional development or integration into physiotherapy curricula. In line with this, the consistently high levels of SDM reported by patients in our study may reflect the effectiveness of advanced practice physiotherapists education in equipping physiotherapists with the clinical skills necessary to support patient involvement in care decisions. Interpreting SDM in physiotherapy remains complex, in part because there is no universally accepted definition or standard for its measurement ^29^. Tools such as CollaboRATE, though widely used for their brevity and patient-centred focus, have been critiqued for potentially oversimplifying the multidimensional nature of SDM, particularly when used as a standalone outcome measure ^18^. This is important context when interpreting our findings, which showed high CollaboRATE scores across both clinical sites. Our results are consistent with our previous research which reported that advanced practice physiotherapists in the UK hold significant knowledge and positive attitudes toward SDM ^26^. However, only a minority reported using structured communication frameworks, and many cited barriers such as, lack of access to decision aids, and restrictive clinical pathways ^26^. These findings raise the possibility that physiotherapists may support the principles of SDM but lack the tools or time to apply them consistently in a structured manner. Further highlighting this gap, another of our recent papers found that SDM levels, measured using OPTION-12 and patient-reported scales, were low in routine consultations without a decision aid, but significantly improved when a decision aid was introduced ^30^. This further supports the view that while physiotherapists may value SDM, structured tools and support systems are necessary to reliably implement it in practice. Importantly, the lack of strong associations between SDM scores and demographic or diagnostic factors suggests that physiotherapists can provide high-quality SDM across diverse populations, even in settings with varying levels of socioeconomic deprivation. This supports the view that SDM is less about patient characteristics and more about clinician behaviour ^31^.

## Strengths and limitations

This study has several strengths, primarily including its large sample size, which to the authors’ knowledge, is by far the largest study within SDM and physiotherapy. The use of both quantitative and qualitative data increases the knowledge and nuance gained from this study. The study was conducted in real-world clinical settings, increasing its clinical applicability to physiotherapy practice. However, several limitations should be acknowledged. We did not collect data on any patients declining participation in the study, and thus it is difficult to rule out whether those who chose not to participate differed systematically from those who did, potentially introducing selection bias. Additionally, while statistical analyses identified small but significant differences in SDM perceptions by site and pain category, the low explanatory power of the regression model suggests that other possible factors, such as consultation length, clinician communication style, or patient health literacy, may play a greater role in SDM experiences ^31^. While our study aimed to examine whether differences in socioeconomic profiles between two locations influenced SDM scores, we found no significant differences. This may suggest that SDM is not primarily driven by patients’ socioeconomic status, but rather by clinicians’ ability to adapt SDM to individual needs. However, due to the lack of individual-level socioeconomic data, we were unable to explore this further. Future research should incorporate such measures to better understand the role of socioeconomic factors in SDM. Because of the nature of our study (including the anonymisation process), we were unable to collect basic demographic information such as age, gender, and pain duration, which limits our ability to characterise the study population and assess whether these factors influenced SDM. A considerable ceiling effect was observed in the CollaboRATE scores, with 95.8% of patients giving the maximum possible score. While this highlights a highly positive perception of SDM in our context, it also limits the ability of the measure to detect variation or change, particularly across subgroups or interventions. Ceiling effects are well-documented in patient-reported experience measures, especially brief instruments like CollaboRATE, and can reduce the sensitivity of statistical analyses to uncover subtle but important differences ^18,20^.

## Conclusion

This study aimed to investigate whether advanced care practitioners succeeded in conducting SDM during musculoskeletal care consultations across two clinical settings and furthermore to qualitatively explore patients’ experiences with advanced practice physiotherapists. Our study shows that patients perceive high levels of SDM in consultations with advanced practice physiotherapists, regardless of clinical site or pain condition. Although statistically significant differences were observed between locations and diagnostic groups, these differences were small, and the explanatory power of associated variables was minimal. The strong ceiling effect observed in CollaboRATE scores highlights both the positive perception of SDM and the limitations of current measurement tools in detecting variability. Importantly, qualitative insights highlighted the central role of clinician-patient interaction, particularly communication, empathy, and professionalism, as key drivers of SDM experiences.

## Supporting information

Credit statement

## Data Availability

All data produced in the present study are available upon reasonable request to the authors

## Declaration of generative AI and AI-assisted technologies in the writing process

During the preparation of this work the author(s) used ChatGPT to support two sections in the introduction for grammatical errors. After using this tool/service, the author(s) reviewed and edited the content as needed and take(s) full responsibility for the content of the publication.

