## Supplementary material for "Advanced Practice Physiotherapists succeed in Shared Decision-making: A Mixed-Methods Study of 5123 Patients with Musculoskeletal Pain": Credit statement

**CRediT author statement: Assessing Shared Decision-Making in Advanced Practice Physiotherapy Care: A Mixed-Methods Study of 5123 Patients with Musculoskeletal Pain**

**Joanne H Thompson**: Conceptualization, Methodology, Formal analysis, Investigation, Data Curation, Writing - Original Draft, Visualization. **Gareth Whelan**: Conceptualization, Methodology, Formal analysis, Investigation, Data Curation, Writing - Review & Editing, Supervision, Project administration. **Glyn Elwyn**: Conceptualization, Methodology, Validation, Formal analysis, Writing - Review & Editing, Supervision**. Kristian Damgaard Lyng**: Conceptualization, Methodology, Software, Validation, Formal analysis, Resources, Data Curation, Writing - Original Draft, Visualization, Supervision, Project administration.
